# Usefulness of quantitative Loop-Mediated Isothermal Amplification Assay in comparison with real time PCR for the diagnosis of invasive pneumococcal disease

**DOI:** 10.1101/2022.12.12.22283380

**Authors:** Sreeram Chandra Murthy Peela, Sujatha Sistla

**Affiliations:** Department of Microbiology, Jawaharlal Institute of Postgraduate Medical Education & Research (JIPMER), Dhanvantri Nagar, Gorimedu, Puducherry – 605006, India

**Author notes:** These authors contributed equally to this work.

**Keywords:** real-time PCR, real-time LAMP, *Streptococcus pneumoniae*, Invasive pneumococcal disease

## Abstract

**Background:** Quantitative LAMP (qLAMP) assay is one of the recent and emerging diagnostic tests for infectious diseases. Only a few studies exist comparing this assay with quantitative real-time PCR (qPCR) for the diagnosis of invasive pneumococcal disease (IPD). The present study was performed to compare the diagnostic performance of qLAMP assay with qPCR targeting autolysin gene for the diagnosis of invasive pneumococcal disease.

**Methodology/Principal Findings:** Ninety six blood samples and 73 CSF samples from patients clinically suspected with community acquired pneumonia and acute meningitis were tested by qPCR and qLAMP assays using previously published primers and protocols. The qPCR was considered as the gold standard test and the diagnostic performance was assessed by calculating sensitivity, specificity, positive and negative predictive values, and kappa coefficient for the level of agreement between the tests. Chi-squared/Fisher exact test was used to compare categorical variables (positive/negative). Thirty two blood samples and 22 CSF samples were positive by qPCR while 24 and 20 samples were positive by qLAMP assay respectively. The sensitivity of qLAMP assay was only 86.4% and 75% when tested on CSF and blood samples respectively. However, the qLAMP assay was in substantial to almost perfect agreement when compared with qPCR. The results were statistically significant in both sample types (P<0.001).

**Conclusions:** The performance of qLAMP assay can vary based on the specimen type. It has very high specificity and had substantial to almost perfect agreement, and thus may be an alternative to qPCR for the diagnosis of IPD.

## Introduction

The Loop Mediated Isothermal Amplification (qLAMP) assay has been used for identification and diagnosis of various pathogens [1–3]. The assay targets six different regions in the primer binding sites, and towards the end of first stage, a stem-loop like structure is formed [1]. The assay uses this loop-like structure for further amplification steps and due to the strand displacement activity of the polymerase used, the assay can proceed isothermally (usually ranging from 60^0^-63^0^C). This advantage circumvents the need for an expensive thermal cycler and the assay can be performed in water bath. To visualize the results, various intercalating dyes or hydroxyl-naphthol blue can be added, or the amplified products can be loaded onto agarose gels where the product shows a ladder-like pattern. The assay can be completed within an hour and around 10^9^ copies of DNA can be produced. It was observed that LAMP assay was more tolerant to various biological substances than PCR. LAMP assay was positive even when 1% serum, plasma or urine was added to the template [4].

Owing to its advantages, it has been tested for diagnosis of pneumococcal meningitis, where a minimum of 10 and 10^4^ copies/reaction of template DNA is required by LAMP and PCR respectively to detect pneumococcus in CSF [5]. The assay was further enhanced by using a loop primer, and results were obtained within 30 minutes [6]. The limit of detection for LAMP assay may vary and it can range from 10 – 90 copies per reaction [5,7]. The limit of detection may vary based on the visualization method or the number of replicates used for each dilution. It was also noted in number of studies targeting pneumococci and other pathogens that performance of LAMP and real-time PCR (qPCR) were comparable [2,7–9].

As evident from many studies, LAMP assay can be a viable alternative for diagnosis of infectious diseases like IPD. It has the potential to be a point-of-care (POC) test and to investigate outbreaks [10]. The present study was thus undertaken to evaluate the usefulness of qLAMP assay for diagnosis of IPD in comparison with qPCR.

## Methods

The study has been approved by the Institute Ethics Committee for Human Studies (JIP/IEC/2015/15/744) and samples collected from May 2015 to August 2018 were included. The samples with sufficient volume of CSF (>0.5ml) and that are negative for any other pathogens by culture were included in the study. Seventy three CSF and 96 blood samples were collected from patients suspected with acute bacterial meningitis (ABM) and community acquired pneumonia (CAP) based on the clinical presentation (S1 Appendix) after obtaining informed consent from the participants or legal guardians. The demographic details are shown in S1 Table. DNA was extracted from these samples using QIAamp DNA Blood Mini kit following manufacturer instructions. The qPCR and qLAMP assays were performed using previously published primers and protocols (S2 Appendix) [6,11]. The 2X real-time PCR master mix (Roche, Basel, Switzerland) and Isothermal Master Mix with intercalating dye (Catalogue number: ISO-001, OptiGene, West Sussex, UK) were used in qPCR and qLAMP assays respectively. Around 2.5μl of DNA was added in qPCR while 5μl was added in qLAMP assay in a total reaction volume of 25μl. The reaction was carried out in ABI QuantStudio 5 (Applied Biosystems, California, USA) by using the standard run settings for TaqMan assay and SYBR-Green assay for qPCR and qLAMP assays respectively. Any sample with C_T_ ≤ 40 was considered as positive in qPCR, while samples with C_T_ ≥ 35 were retested. Similar to qPCR, the time to detection for amplification was noted as the cycle threshold (C_LAMP_) values for qLAMP assay. Both C_T_ and C_LAMP_ were expressed as median with interquartile ranges (IQR). The results by qPCR were considered as true positive. Diagnostic performance of qLAMP assay was estimated by calculating sensitivity, specificity, positive and negative predictive values. The agreement between two assays was quantified using Cohen’s kappa coefficient and interpreted accordingly. All the analysis was carried out in OpenEpi v3.01.

## Results

Twenty CSF and 24 blood samples were identified to be positive (Table 1) by qLAMP assay, while 22 and 32 specimens were positive by qPCR. The median C_T_ values were similar for both meningitis (28.7; IQR=22.8-33.1) and pneumonia group (28.4; IQR=20.9-30.6) while median C_LAMP_ values varied slightly (11.9 and IQR=10.5-13.7 and 11 and IQR=8.4-11.8 respectively) (S2 Table). One CSF specimen negative by qPCR was positive in the qLAMP assay (C_LAMP_ = 18.2), while eight blood samples and three CSF samples positive by qPCR were negative by qLAMP assay. The median C_T_ values were higher (37.9; IQR=35.7-38.3) in samples negative by qLAMP assay than those positive by it (24; IQR=18.2-28.8) when both pneumonia and meningitis groups were analyzed together (S2 Table). The overall sensitivity of qLAMP was slightly higher while testing CSF specimens, and for both the specimen types (Table 2), the two assays had substantial to almost perfect agreement (kappa coefficient > 0.8) (Table 1).

**Table 1:** Comparison of results between real-time PCR (qPCR) and real-time LAMP (qLAMP) assays in blood and cerebrospinal fluid (CSF) samples. 95% Confidence intervals for Kappa coefficient were shown in parentheses

| qPCR |  |  |  |  | Kappa coefficient | P-value |
| --- | --- | --- | --- | --- | --- | --- |
|  |  |  | Positive | Negative |  |  |
| Blood | qLAMP | Positive | 24 | 0 | 0.8(0.604,0.996) | <0.001 |
|  |  | Negative | 8 | 64 |  |  |
|  |  | Total | 32 | 64 |  |  |
| CSF | qLAMP | Positive | 19 | 1 | 0.899(0.67,1.13) | <0.001 |
|  |  | Negative | 3 | 50 |  |  |
|  |  | **Total** | 22 | 51 |  |  |

**Table 2:** Diagnostic metrics of real-time LAMP (qLAMP) assays in blood and cerebrospinal fluid (CSF) samples. 95% Confidence intervals were shown in parentheses *Positive Predictive Value ^#^Negative Predictive Value

| **qLAMP** | **Sensitivity** | **Specificity** | **PPV\*** | **NPV#** |
| --- | --- | --- | --- | --- |
| CSF | 86.4(66.7,95.3) | 98(88.7,99.7) | 95(73.4,99.1) | 94.3(84.6,98.1) |
| Blood | 75(57.9,86.6) | 100(94.3,100) | 100(86.2,100) | 88.9(79.6,94.3) |

## Discussion

The use of qLAMP assay diagnosis of pneumococcal meningitis as a sensitive and rapid method was described by Kim *et al* [6]. In the present study a closed tube technique for interpretation of the test result was employed to prevent contamination through aerosols. To test the utility of qLAMP assay, all the samples that were positive by qPCR were included along with those negative for pneumococcus. Three CSF and eight blood specimens positive by qPCR were qLAMP-negative. One of the explanations for these qLAMP-negative specimens may be the time lag between the performances of both the molecular assays, where qLAMP was performed after one year on DNA stored at −80^0^C. Furthermore, all the specimens negative by qLAMP assay had C_T_ > 35, implying a very low concentration of DNA which may have undergone degradation during the prolonged storage. However, qLAMP assay can detect a few additional cases when tested on CSF specimens (one qPCR negative specimen was positive by qLAMP assay in the present study). The same was true in another study where an additional positive result in qLAMP assay was observed [7].

For the detection of pneumococcal meningitis, LAMP assay had 100% sensitivity and 83% specificity when compared with PCR [6]. Furthermore, the clinical specificity and negative predictive value (NPV) for LAMP assay while testing on CSF samples was found to be higher. In another study, both the assays showed significant similarity, with 100% sensitivity and 99.3% specificity when testing on multiple specimens from IPD patients [7]. Similar to this finding, in our study the specificity of LAMP assay was 98%, but NPV was only 94.3%. Interestingly, the PPV was 100% for the blood samples and 95% for CSF samples in the present study. While the sensitivity of qLAMP assay was low, the specificity was at par with that reported previously.

A few studies found that real-time PCR is more sensitive than qLAMP assay in detecting pneumococci and other microbial pathogens. In a recent study by Hector et al, the limit of detection between qPCR and qLAMP assay varied slightly (90 copies/reaction for qLAMP and 20 copies/reaction for qPCR) [7]. Wang *et al* found that the qLAMP assay had a sensitivity of only 89.1% while that of real-time PCR was 91.3% while detecting orf virus [8]. Lin *et al* in their study found that the limit of detection of qLAMP assay was higher than that of real-time PCR (10 fg/μl and 1 fg/μl respectively) for detection of Toxoplasma in blood [9]. Chen *et al* on the other hand found that the qLAMP assay and real-time PCR had similar limits of detection while targeting Japanese Encephalitis virus [2]. Nevertheless in all these studies, the results of qLAMP assay were in excellent agreement with that of real-time PCR, and hence qLAMP assay can be a viable alternative.

The limitations in the present study were: a time gap of one year between performing qLAMP and qPCR assays, and limit of detection for these assays was not calculated. Nonetheless this study was one of the few reports that tried to compare established qPCR assay with the emerging qLAMP assay and testing for both blood and CSF specimens which are frequently tested for the diagnosis of IPD.

## Conclusions

The performance of qLAMP assay can vary based on the specimen type used for testing. Owing to its substantial agreement and high specificity when compared with qPCR, it may be considered an alternative test for diagnosis of pneumococcal meningitis and bacteremia. However in samples with low bacterial load, qLAMP assay may produce false negative results.

## Data Availability

The data is presented in supplementary material. Full data can be available upon requesting the corresponding author

## Supporting information

**S1 Appendix:** Criteria for clinical diagnosis

**S2 Appendix:** Protocols. *From a stock of 10 μM; ^#^From a stock of 50 μM

**S1 Table:** Demographic details. Frequencies and percentages are shown for each group

**S2 Table:** Cycle threshold values. Zero indicates a negative result. C_T_ – cycle threshold in qPCR; C_LAMP_ – cycle threshold in qLAMP. Any sample with C_T_ >35 were retested (duplicates) and mean values are shown

## Notes

### Competing Interest Statement

The authors have declared no competing interest.

### Funding Statement

JIPMER Intramural Research Fund - JIP/RES/INTRA-PHD/PHS1/01/2016-17

### Author Declarations

The study has been approved by the Institute Ethics Committee for Human Studies (JIP/IEC/2015/15/744)

